# Increased disparity in routine infant vaccination during COVID-19

**DOI:** 10.1101/2022.06.30.22277115

**Authors:** Christiaan H. Righolt, Gupreet Pabla, Salaheddin M. Mahmud

## Abstract

**Background:** COVID-19 restrictions and its impact on healthcare resources have reduced routine infant vaccine uptake, although some report that this effect was short-lived. These prior studies mostly described entire populations, but disparities in uptake may have changed during the pandemic due to differential access to healthcare.

**Objectives:** We aimed to examine disparities in the reduction in routine infant vaccine uptake during the COVID-19 pandemic in Manitoba, Canada.

**Methods:** We assessed vaccine uptake for routine infant vaccines for a pre-pandemic and pandemic subcohort. We assessed how the reduction in vaccine uptake differed by gender, neighborhood income quintile and region of residence. For each evaluation age, we limited the pandemic subcohort to children reaching this milestone age on/before November 30, 2021.

**Results:** Vaccine uptake was about 5-10% lower during the pandemic. The groups most vulnerable to COVID-19 saw the largest reductions in vaccine uptake, with an ongoing downward trend throughout the pandemic. Children in the lowest income neighborhoods saw a 17% reduction in diphtheria, tetanus, and acellular pertussis dose 4 uptake at 24 months, 4.4-fold that of high-income neighborhoods, and an 11% reduction in measles, mumps, rubella (MMR) vaccine uptake at 24 months, 5.6-fold that of high-income neighborhoods. The largest reductions were for low-income Northern residents and smallest for high-income Winnipeg residents, e.g. 16-fold larger for MMR at 24 months (79:94 pre-pandemic to 65:93 during the pandemic).

**Conclusions:** While privileged children have similar high vaccine uptake as before the pandemic, children in populations hardest hit by COVID-19 continue seeing concerning reductions in routine infant vaccination. It is imperative that infant vaccination rates are increased, especially in communities with lower socioeconomic status, as a failure to do so could lead to persistent rebound epidemics in the most vulnerable populations.

**Synopsis:** *Study question:* How did COVID-19 and its restrictions affect routine infant vaccine uptake?

*What’s already known:* We know that vaccine uptake in infants decreased during the pandemic. We do not know whether this affected everyone equally or whether the pandemic worsened existing disparities in vaccine uptake.

*What this study adds:* Although vaccine uptake was not affected in wealthy urban neighborhoods, the reduction in uptake was largest, and continued on a downward trend, for groups with the lowest baseline vaccine uptake. Only two-thirds of children, instead of the 4/5th before the pandemic, in the remote, predominantly Indigenous Northern region received a measles vaccine by their second birthday.

## Introduction

COVID-19 restrictions and its impact on healthcare resources have reduced routine infant vaccine uptake,^1,2^ although some report that this effect was short-lived.^3^ Many studies examined the population as a whole, but disparities (by socioeconomic status) in uptake may have changed during the pandemic due to differential access to healthcare. We examined the disparity and reduction of these vaccinations in Manitoba, a Canadian province with 1.3 million residents.

## Methods

Manitoba Health (MH) is the publicly funded health insurance agency providing comprehensive health insurance to the province’s residents. Coverage is universal, with no eligibility distinction based on age or income, and participation rates are very high (>99%).^4^ Insured services include hospital, physician and preventive services including vaccinations. MH maintains several centralized, administrative electronic databases that are linkable using a unique personal health identification number (PHIN). The completeness and accuracy of these databases are well established.^5,6^

The MH Population Registry (MHPR) tracks addresses and dates of birth, death and health insurance coverage for all insured persons. The Manitoba Immunization Monitoring System (MIMS) and its successor the Public Health Information Management System (PHIMS) is the population-based province-wide registry that contains records of all childhood vaccinations administered in Manitoba since 1988 and all adult vaccinations since 2000.^7^ Infant vaccination is primarily performed by physicians in Winnipeg, the main urban center in the province housing over half the population, and by public health nurses outside of Winnipeg. Information, including vaccine type and administration date, is captured either through direct data entry (for vaccines administered by public health staff) or through physician claims. Validation studies have shown high completeness and accuracy of MIMS data, with less than 2% of vaccinations coded incorrectly, and accurate service dates recorded 98% of the time.^7^

We identified a *study cohort* of all children born in Manitoba between January 2017 and November 2020 from the MHPR. We split this study cohort into 3 subcohorts based on the start of the COVID-19 related restrictions in Manitoba on March 15, 2020 as well as the vaccine recommendation age and the evaluation age at which vaccine uptake was determined (Figure 1). The *pre-pandemic subcohort* consisted of children who reached the evaluation age before the start of restrictions. The *pandemic subcohort* consisted of children who reached the vaccine recommendation age after the start of restrictions. The *intermediate subcohort* consisted of children born between these subcohorts.

**Figure 1.**
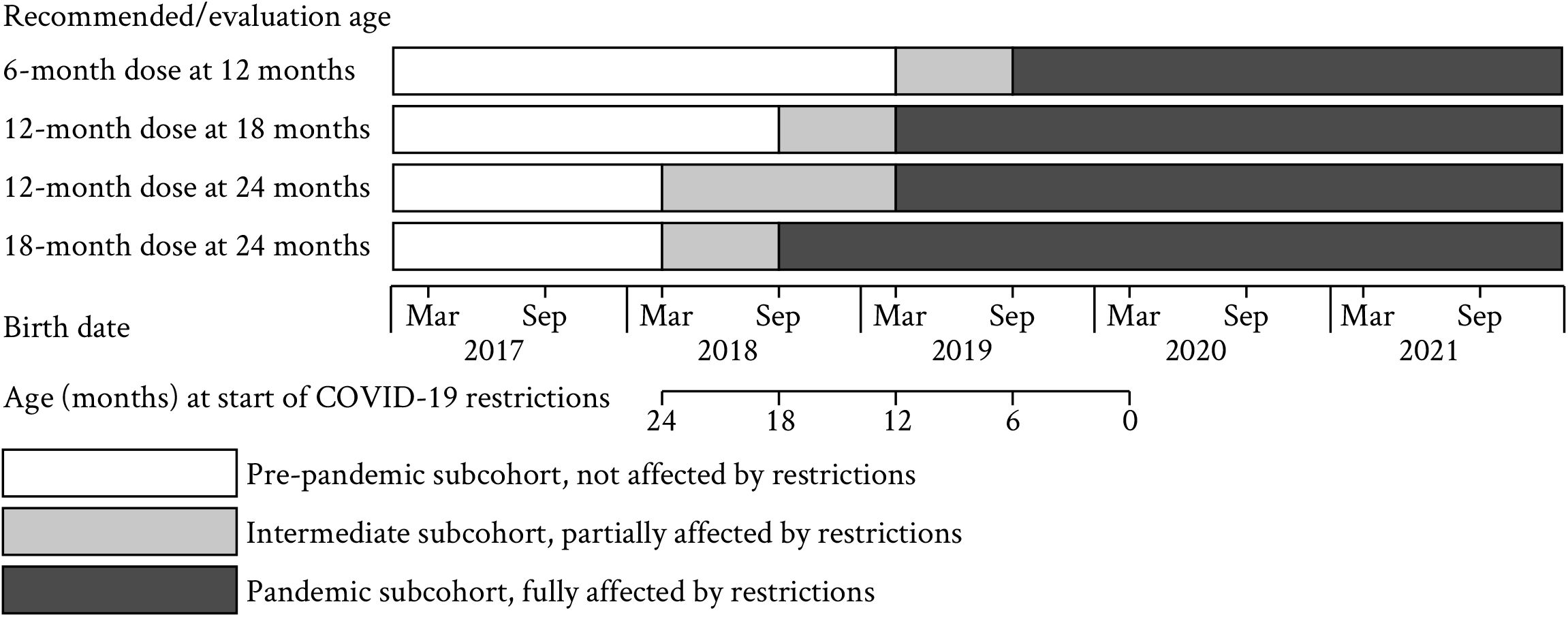
Study cohort and subcohorts based on the start of the COVID-19 related restriction in Manitoba (on March 15, 2020), the age at which the vaccine is recommended (Table S1), and the evaluation age for vaccine uptake.

We linked these subcohorts to MIMS/PHIMS to assess vaccine uptake for routine infant vaccines (Table S1). We assessed how the reduction in vaccine uptake differed by gender, neighborhood income quintile (obtained from the 2016 Canadian census) and region of residence. For each evaluation age, we limited the pandemic subcohort to children who reached this evaluation age (12, 18, or 24 months) on/before November 30, 2021 and who resided in Manitoba from birth to the evaluation age to ensure their vaccination record was complete.

## Results

Vaccine uptake was about 5-10% lower during the pandemic (Table 1, Table S2), affecting boys and girls similarly. Uptake in the intermediate subcohort was mostly similar to the pre-pandemic subcohort (Table S3). The groups most vulnerable to COVID-19^8^ saw the largest reductions in vaccine uptake, with an ongoing downward trend throughout the pandemic (Figure 2). Children in the lowest income neighborhoods saw a 17% reduction in diphtheria, tetanus, and acellular pertussis (DTaP) dose 4 uptake at 24 months, 4.4-fold that of high-income neighborhoods, and an 11% reduction in measles, mumps, rubella (MMR) vaccine uptake at 24 months, 5.6-fold that of high-income neighborhoods. Uptake reductions were largest in the ∼75% Indigenous Northern region: 6-fold that of Winnipeg, which houses >50% of Manitobans, for MMR at 24 months.

**Table 1.** Changes in vaccine uptake (%) of routine childhood vaccines during the pandemic (data ending November 30, 2021).

| DTaP | Dose 3 uptake at 12 months<sup>a</sup> |  |  |  | Dose 4 uptake at 24 months<sup>b</sup> |  |  |  |
| --- | --- | --- | --- | --- | --- | --- | --- | --- |
|  | Pre-pandemic | Pandemic | Relative change | Fold change | Pre-pandemic | Pandemic | Relative change | Fold change |
| Overall | 80 | 74 | -7% |  | 71 | 64 | -10% |  |
| Gender |  |  |  |  |  |  |  |  |
| Male | 80 | 74 | -8% | ref. | 71 | 64 | -10% | ref. |
| Female | 79 | 74 | -7% | 0.9 | 71 | 64 | -10% | 1.0 |
| Income quintile |  |  |  |  |  |  |  |  |
| Q1 (lowest) | 71 | 62 | -13% | 6.1 | 61 | 50 | -17% | 4.4 |
| Q2 | 80 | 72 | -10% | 4.6 | 71 | 61 | -14% | 3.5 |
| Q3 | 83 | 79 | -5% | 2.6 | 75 | 71 | -6% | 1.5 |
| Q4 | 82 | 77 | -7% | 3.2 | 75 | 70 | -8% | 1.9 |
| Q5 (highest) | 85 | 84 | -2% | ref. | 79 | 76 | -4% | ref. |
| Unknown | 81 | 81 | 1% | -0.2 | 70 | 71 | 2% | -0.4 |
| Residence |  |  |  |  |  |  |  |  |
| Rural | 70 | 60 | -14% | 5.5 | 62 | 53 | -15% | 2.3 |
| Urban | 87 | 85 | -3% | ref. | 79 | 74 | -6% | ref. |
| Unknown | 75 | 67 | -12% | 4.5 | 55 | 42 | -23% | 3.6 |
| Regional health authority of residence |  |  |  |  |  |  |  |  |
| Winnipeg | 87 | 85 | -3% | ref. | 79 | 74 | -6% | ref. |
| Interlake-Eastern | 78 | 65 | -16% | 6.6 | 68 | 60 | -11% | 1.8 |
| Northern | 64 | 50 | -22% | 8.6 | 51 | 39 | -25% | 4.0 |
| Southern | 66 | 58 | -12% | 4.6 | 60 | 51 | -15% | 2.4 |
| Prairie Mountain | 82 | 77 | -7% | 2.6 | 74 | 68 | -9% | 1.4 |
| Public Trustee / In CFS care | 75 | 67 | -12% | 4.6 | 55 | 42 | -23% | 3.6 |
| MMR | Dose 1 uptake at 18 months<sup>c</sup> |  |  |  | Dose 1 uptake at 24 months<sup>d</sup> |  |  |  |
|  | Pre-pandemic | Pandemic | Relative change | Fold change | Pre-pandemic | Pandemic | Relative change | Fold change |
| Overall | 82 | 75 | -9% |  | 86 | 80 | -7% |  |
| Gender |  |  |  |  |  |  |  |  |
| Male | 82 | 75 | -9% | ref. | 86 | 80 | -7% | ref. |
| Female | 82 | 76 | -8% | 0.9 | 86 | 80 | -7% | 1.1 |
| Income quintile |  |  |  |  |  |  |  |  |
| Q1 (lowest) | 76 | 66 | -14% | 3.5 | 83 | 73 | -11% | 5.6 |
| Q2 | 83 | 74 | -12% | 3.0 | 88 | 80 | -10% | 4.7 |
| Q3 | 84 | 79 | -6% | 1.5 | 87 | 84 | -3% | 1.5 |
| Q4 | 83 | 77 | -7% | 1.9 | 87 | 81 | -7% | 3.3 |
| Q5 (highest) | 87 | 84 | -4% | ref. | 89 | 87 | -2% | ref. |
| Unknown | 84 | 82 | -2% | 0.5 | 87 | 86 | -1% | 0.5 |
| Residence |  |  |  |  |  |  |  |  |
| Rural | 75 | 64 | -15% | 3.6 | 80 | 71 | -12% | 4.0 |
| Urban | 89 | 85 | -4% | ref. | 92 | 89 | -3% | ref. |
| Unknown | 80 | 60 | -25% | 6.1 | 81 | 53 | -34% | 11.5 |
| Regional health authority of residence |  |  |  |  |  |  |  |  |
| Winnipeg | 89 | 85 | -4% | ref. | 92 | 89 | -3% | ref. |
| Interlake-Eastern | 80 | 70 | -12% | 3.1 | 83 | 78 | -6% | 2.3 |
| Northern | 76 | 57 | -24% | 6.3 | 85 | 71 | -16% | 6.0 |
| Southern | 68 | 60 | -12% | 3.2 | 73 | 64 | -12% | 4.5 |
| Prairie Mountain | 84 | 76 | -10% | 2.5 | 88 | 80 | -9% | 3.3 |
| Public Trustee / In CFS care | 80 | 60 | -25% | 6.5 | 81 | 53 | -34% | 12.4 |

DTaP = Diphtheria, tetanus, and acellular pertussis vaccine
MMR = Measles, mumps, and rubella vaccine
<sup>a</sup> Recommended age is 6 months; Pre-pandemic cohort, born January 1, 2017 - March 15, 2019; Pandemic cohort, born September 15, 2019 and later
<sup>b</sup> Recommended age is 18 months; Pre-pandemic cohort, born January 1, 2017 - March 15, 2018; Pandemic cohort, born September 15, 2018 and later
<sup>c</sup> Recommended age is 12 months; Pre-pandemic cohort, born January 1, 2017 - September 15, 2018; Pandemic cohort, born March 15, 2019 and later
<sup>d</sup> Recommended age is 12 months; Pre-pandemic cohort, born January 1, 2017 - March 15, 2018; Pandemic cohort, born March 15, 2019 and later

**Figure 2.**
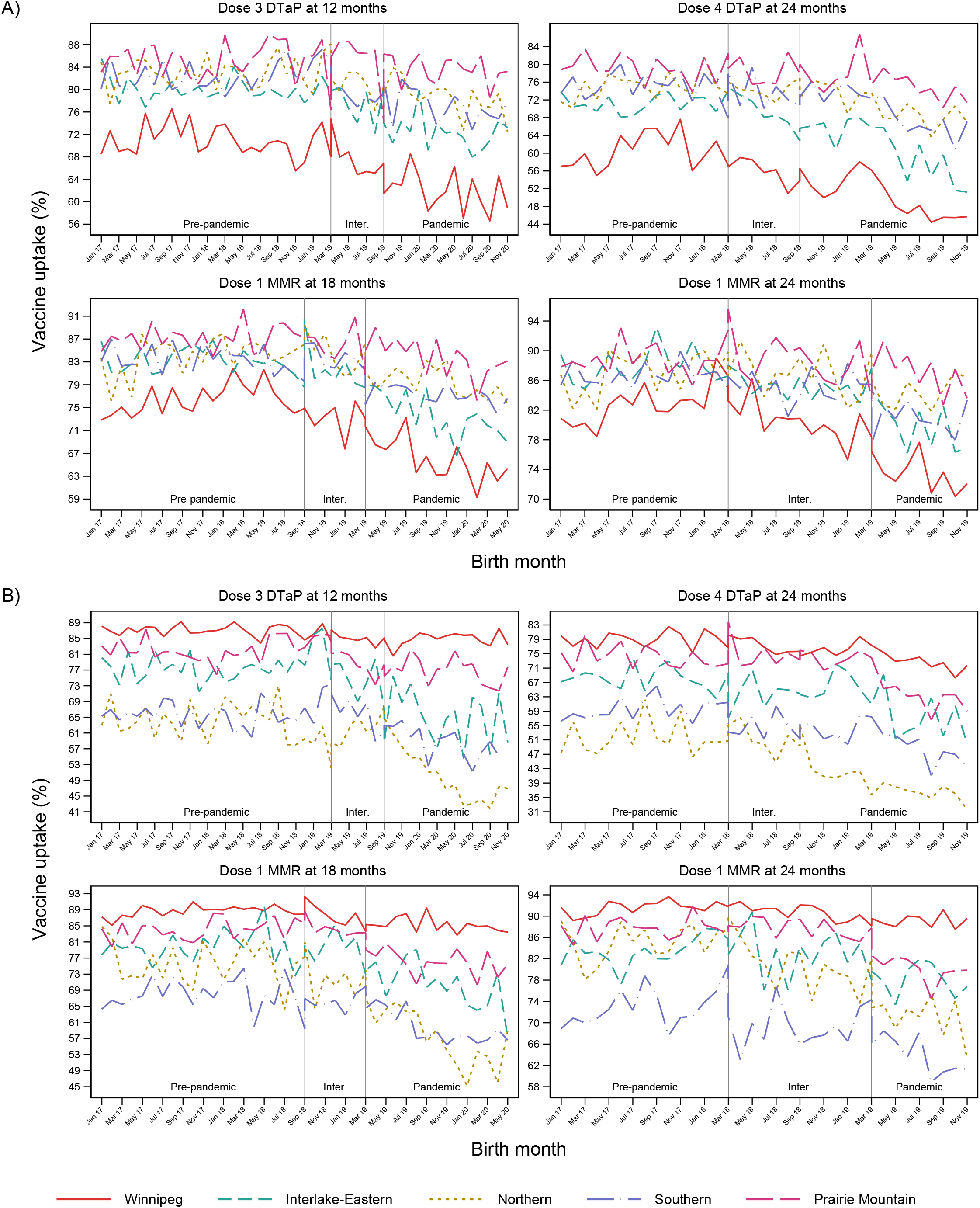
Vaccine uptake (percentage of the population) of DTaP (diphtheria, tetanus, and acellular pertussis) dose 3 at 12 months and dose 4 at 24 months, and MMR (measles, mumps, rubella) dose 1 at 18 and 24 months by neighborhood income quintile (A) and regional health authority (B) according to month of birth. Winnipeg is the main urban center of Manitoba, housing >50% of the provincial population; Northern Manitoba is a remote, predominantly (∼75%) Indigenous region.

Disparity by income increased in each region (Table S4): The lower the neighborhood income quintile, the higher was the reduction in vaccine uptake. Regions with lower pre-pandemic vaccine uptake saw the largest reductions during the pandemic. The largest reductions were for low-income Northern residents and smallest for high-income Winnipeg residents, e.g., 16-fold larger for MMR at 24 months (79:94 pre-pandemic to 65:93 during the pandemic; Table S4). Although reductions were small and rebounded in Winnipeg and high-income neighborhoods (Figure 2), low-income neighborhoods and other regions, most notably the Northern region, were on a continuous downward trend in vaccine uptake. These trends held across all vaccines.

## Discussion

Reductions in routine infant vaccine uptake were largest, and continued on a downward trend, for groups with the lowest baseline vaccine uptake. COVID-19-related reassignment of public health nurses, who provide infant vaccination outside of Winnipeg, may explain some of these differences in vaccine uptake. In Quebec, overall vaccination rates rebounded quickly after an initial reduction;^3^ in Ontario, vaccine uptake reduced and remained lower during the pandemic.^9^ Neither of these other studies explicitly accounted for the intermediate subcohort or showed the precipitous and continuous decline we observed for the most vulnerable infants. A Louisiana study found that disadvantaged children (Medicaid beneficiaries in low-income households) had a 30% drop in vaccinations during the initial COVID-19 restrictions,^10^ but did not include a comparison with children in high-income households to compare with our observed increase in disparity. As other jurisdictions face similar disparities in access to healthcare,^11^ it is likely that similar differential reductions in vaccine uptake occurred elsewhere.

Confounding is unlikely to bias these results, as most children in this analysis were conceived before the first case of COVID-19 in Manitoba. Any potential (differential) changes in birth rates due to COVID-19 and its restrictions would occur for births after 2020. This study is limited by the completeness and timeliness of this data, although the quality of these databases are well-established.^5^

In conclusion, while privileged children have similar high vaccine uptake as before the pandemic, children in populations hardest hit by COVID-19 continue seeing concerning reductions in routine infant vaccination. Because current vaccine uptake is lower than the herd immunity threshold for many vaccine-preventable diseases, it is imperative that infant vaccination rates are increased, especially in communities with lower socioeconomic status, as a failure to do so could lead to persistent rebound epidemics in the most vulnerable populations.

## Data Availability

Data used in this article was derived from administrative health and social data as a secondary use. The data was provided under specific data sharing agreements only for approved use at the Manitoba Centre for Health Policy (MCHP). The original source data is not owned by the researchers or MCHP and as such cannot be provided to a public repository. The original data source and approval for use has been noted in the acknowledgments of the article. Where necessary, source data specific to this article or project may be reviewed at MCHP with the consent of the original data providers, along with the required privacy and ethical review bodies.

## Abbreviations

COVID-19: coronavirus disease 2019;
DTaP: Diphtheria, tetanus, and acellular pertussis;
MH: Manitoba Health;
MMR: measles, mumps, rubella

## Ethics approval

This study was approved by the University of Manitoba Research Ethics Board and by the Manitoba Health Health Information Privacy Committee.

## Acknowledgements

The authors acknowledge the Manitoba Centre for Health Policy for use of data contained in the Manitoba Population Research Data Repository under project #2021-015 (HIPC #2020/2021–73, REB # HS24593(H2021:026). The results and conclusions are those of the authors and no official endorsement by the Manitoba Centre for Health Policy or Manitoba Health is intended or should be inferred. Data used in this study are from the Manitoba Population Research Data Repository housed at the Manitoba Centre for Health Policy, University of Manitoba and were derived from data provided by Manitoba Health.

## Author contributions

CHR conceptualized and designed the study, oversaw the analysis, drafted the initial manuscript, and reviewed and revised the manuscript. GP performed the analysis, and reviewed and revised the manuscript. SMM conceptualized and designed the study, and reviewed and revised the manuscript.

