## Supplement for "Increased disparity in routine infant vaccination during COVID-19"

**Supplementary tables**

Table S1 **Recommended infant vaccination schedule in Manitoba**

| 2 months | 4 months | 6 months | 12 months | 18 months |
|---|---|---|---|---|
| DTaP | DTaP | DTaP | MMR | DTaP |
| Polio | Polio | HiB | Varicella | Polio |
| HiB | HiB | Rotavirus[1] | PCV | Hib |
| PCV | PCV | | MenC-C | |
| Rotavirus | Rotavirus | | | |

DTaP = Diphtheria, tetanus, and acellular pertussis vaccine

HiB = Haemophilus influenzae type b vaccine

MenC-C = Meningococcal conjugate serogroup C vaccine

MMR = Measles, mumps, and rubella vaccine

PCV = 13-valent pneumococcal conjugate vaccine

Note that some of these are administered in combination as one product, e.g., MMR and varicella.

[1]Only for kids born March 2018 and later. This vaccine is excluded from the analysis, because the 6-month dose was introduced shortly before the start of the pandemic.

Table S2 **Changes in vaccine uptake (%) of routine childhood vaccines during the pandemic (data ending November 30, 2021).**

| **Polio** | **Dose 2 uptake at 12 months[a]** | | | |
|---|---|---|---|---|
| | **Pre-pandemic** | **Pandemic** | **Relative change** | **Fold change** |
| Overall | 88 | 83 | -6% | |
| Gender | | | | |
| Male | 88 | 82 | -7% | ref. |
| Female | 88 | 83 | -6% | 0.8 |
| Income quintile | | | | |
| Q1 (lowest) | 85 | 77 | -9% | 2.9 |
| Q2 | 89 | 83 | -7% | 2.2 |
| Q3 | 90 | 85 | -5% | 1.5 |
| Q4 | 88 | 83 | -6% | 1.7 |
| Q5 (highest) | 91 | 88 | -3% | ref. |
| Unknown | 89 | 86 | -3% | 1.0 |
| Residence | | | | |
| Rural | 81 | 71 | -12% | 6.0 |
| Urban | 94 | 92 | -2% | ref. |
| Unknown | 88 | 100 | 14% | -7.0 |
| Winnipeg region of residence | | | | |
| Northern suburbs | 95 | 92 | -3% | 2.1 |
| Inner city | 91 | 88 | -3% | 2.3 |
| Southern suburbs | 95 | 94 | -1% | ref. |
| Regional health authority of residence | | | | |
| Winnipeg | 94 | 92 | -2% | ref. |
| Interlake-Eastern | 86 | 76 | -13% | 6.1 |
| Northern | 84 | 72 | -15% | 7.2 |
| Southern | 73 | 65 | -12% | 5.9 |
| Prairie Mountain | 89 | 85 | -5% | 2.3 |
| Public Trustee / In CFS care | 88 | 100 | 14% | -6.8 |
| **PCV** | **Dose 3 uptake at 18 months[b]** | | | |
| | **Pre-pandemic** | **Pandemic** | **Relative change** | **Fold change** |
| Overall | 78 | 72 | -8% | |
| Gender | | | | |
| Male | 79 | 72 | -8% | ref. |

| | | | | |
|---|---|---|---|---|
| Female | 78 | 73 | -7% | 0.9 |
| Income quintile | | | | |
| Q1 (lowest) | 71 | 64 | -11% | 2.9 |
| Q2 | 80 | 72 | -10% | 2.7 |
| Q3 | 81 | 76 | -7% | 1.8 |
| Q4 | 80 | 74 | -7% | 1.9 |
| Q5 (highest) | 84 | 80 | -4% | ref. |
| Unknown | 81 | 78 | -3% | 0.7 |
| Residence | | | | |
| Rural | 70 | 63 | -10% | 1.7 |
| Urban | 85 | 80 | -6% | ref. |
| Unknown | 78 | 55 | -29% | 4.9 |
| Winnipeg region of residence | | | | |
| Northern suburbs | 86 | 81 | -5% | 0.9 |
| Inner city | 79 | 73 | -8% | 1.4 |
| Southern suburbs | 87 | 82 | -6% | ref. |
| Regional health authority of residence | | | | |
| Winnipeg | 85 | 80 | -6% | ref. |
| Interlake-Eastern | 76 | 68 | -10% | 1.7 |
| Northern | 70 | 61 | -13% | 2.2 |
| Southern | 64 | 58 | -9% | 1.6 |
| Prairie Mountain | 80 | 73 | -9% | 1.5 |
| Public Trustee / In CFS care | 78 | 55 | -29% | 5.1 |

| **MenC-C** | **Dose 1 uptake at 24 months[c]** | | | |
|---|---|---|---|---|
| | **Pre-pandemic** | **Pandemic** | **Relative change** | **Fold change** |
| Overall | 86 | 80 | -7% | |
| Gender | | | | |
| Male | 86 | 80 | -7% | ref. |
| Female | 86 | 80 | -7% | 1.1 |
| Income quintile | | | | |
| Q1 (lowest) | 83 | 73 | -11% | 4.8 |
| Q2 | 88 | 79 | -9% | 4.0 |
| Q3 | 87 | 84 | -3% | 1.2 |
| Q4 | 87 | 81 | -7% | 2.8 |
| Q5 (highest) | 89 | 87 | -2% | ref. |
| Unknown | 87 | 86 | -1% | 0.6 |
| Residence | | | | |

| | | | | |
|---|---|---|---|---|
| Rural | 80 | 70 | -12% | 4.1 |
| Urban | 92 | 89 | -3% | ref. |
| Unknown | 77 | 53 | -31% | 10.9 |
| Winnipeg region of residence | | | | |
| Northern suburbs | 93 | 90 | -3% | 2.0 |
| Inner city | 87 | 83 | -5% | 3.6 |
| Southern suburbs | 93 | 92 | -1% | ref. |
| Regional health authority of residence | | | | |
| Winnipeg | 92 | 89 | -3% | ref. |
| Interlake-Eastern | 83 | 78 | -6% | 2.2 |
| Northern | 85 | 71 | -16% | 6.3 |
| Southern | 72 | 63 | -12% | 4.6 |
| Prairie Mountain | 87 | 79 | -10% | 3.7 |
| Public Trustee / In CFS care | 77 | 53 | -31% | 12.0 |
| **Varicella** | **Dose 1 uptake at 24 months[c]** | | | |
| | **Pre-pandemic** | **Pandemic** | **Relative change** | **Fold change** |
| Overall | 86 | 80 | -7% | |
| Gender | | | | |
| Male | 85 | 80 | -6% | ref. |
| Female | 86 | 80 | -7% | 1.1 |
| Income quintile | | | | |
| Q1 (lowest) | 82 | 73 | -11% | 5.1 |
| Q2 | 87 | 79 | -9% | 4.2 |
| Q3 | 86 | 84 | -3% | 1.2 |
| Q4 | 86 | 80 | -6% | 2.8 |
| Q5 (highest) | 88 | 86 | -2% | ref. |
| Unknown | 85 | 85 | 0% | -0.1 |
| Residence | | | | |
| Rural | 79 | 70 | -11% | 4.1 |
| Urban | 91 | 88 | -3% | ref. |
| Unknown | 81 | 53 | -34% | 12.0 |
| Winnipeg region of residence | | | | |
| Northern suburbs | 92 | 89 | -3% | 3.0 |
| Inner city | 87 | 82 | -6% | 5.5 |
| Southern suburbs | 92 | 91 | -1% | ref. |
| Regional health authority of residence | | | | |
| Winnipeg | 91 | 89 | -3% | ref. |

| | | | | |
|---|---|---|---|---|
| Interlake-Eastern | 83 | 77 | -7% | 2.6 |
| Northern | 85 | 71 | -16% | 6.3 |
| Southern | 71 | 63 | -11% | 4.4 |
| Prairie Mountain | 88 | 80 | -9% | 3.4 |
| Public Trustee / In CFS care | 81 | 53 | -34% | 13.1 |

| **Polio** | **Dose 3 uptake at 24 months[d]** | | | |
|---|---|---|---|---|
| | **Pre-pandemic** | **Pandemic** | **Relative change** | **Fold change** |
| Overall | 87 | 84 | -3% | |
| Gender | | | | |
| Male | 87 | 84 | -3% | ref. |
| Female | 87 | 84 | -4% | 1.2 |
| Income quintile | | | | |
| Q1 (lowest) | 83 | 78 | -6% | 6.7 |
| Q2 | 88 | 85 | -4% | 4.6 |
| Q3 | 88 | 87 | -2% | 1.9 |
| Q4 | 87 | 85 | -2% | 2.2 |
| Q5 (highest) | 90 | 89 | -1% | ref. |
| Unknown | 89 | 88 | -1% | 1.2 |
| Residence | | | | |
| Rural | 80 | 76 | -5% | 3.4 |
| Urban | 93 | 91 | -2% | ref. |
| Unknown | 84 | 85 | 1% | -0.6 |
| Winnipeg region of residence | | | | |
| Northern suburbs | 94 | 92 | -2% | 1.7 |
| Inner city | 90 | 86 | -4% | 4.1 |
| Southern suburbs | 94 | 93 | -1% | ref. |
| Regional health authority of residence | | | | |
| Winnipeg | 93 | 92 | -2% | ref. |
| Interlake-Eastern | 85 | 82 | -3% | 1.9 |
| Northern | 83 | 75 | -10% | 5.5 |
| Southern | 73 | 70 | -5% | 3.0 |
| Prairie Mountain | 88 | 86 | -2% | 1.0 |
| Public Trustee / In CFS care | 84 | 85 | 1% | -0.5 |

| **HiB** | **Dose 4 uptake at 24 months[d]** | | | |
|---|---|---|---|---|
| | **Pre-pandemic** | **Pandemic** | **Relative change** | **Fold change** |
| Overall | 71 | 63 | -10% | |
| Gender | | | | |

| | | | | |
|---|---|---|---|---|
| Male | 70 | 63 | -10% | ref. |
| Female | 71 | 63 | -10% | 1.1 |
| Income quintile | | | | |
| Q1 (lowest) | 60 | 49 | -18% | 3.9 |
| Q2 | 71 | 60 | -14% | 3.1 |
| Q3 | 75 | 70 | -6% | 1.3 |
| Q4 | 75 | 69 | -8% | 1.7 |
| Q5 (highest) | 78 | 75 | -5% | ref. |
| Unknown | 70 | 71 | 2% | -0.4 |
| Residence | | | | |
| Rural | 61 | 52 | -15% | 2.2 |
| Urban | 78 | 73 | -7% | ref. |
| Unknown | 55 | 42 | -23% | 3.4 |
| Winnipeg region of residence | | | | |
| Northern suburbs | 81 | 75 | -7% | 1.5 |
| Inner city | 69 | 60 | -13% | 2.9 |
| Southern suburbs | 81 | 77 | -5% | ref. |
| Regional health authority of residence | | | | |
| Winnipeg | 78 | 73 | -7% | ref. |
| Interlake-Eastern | 68 | 60 | -12% | 1.7 |
| Northern | 51 | 38 | -25% | 3.7 |
| Southern | 59 | 50 | -16% | 2.4 |
| Prairie Mountain | 74 | 67 | -9% | 1.4 |
| Public Trustee / In CFS care | 55 | 42 | -23% | 3.4 |

PCV = 13-valent pneumococcal conjugate vaccine

MenC-C = Meningococcal conjugate serogroup C vaccine

HiB =Haemophilus influenzae type b vaccine

[a] Recommended age is 4 months; Pre-pandemic cohort, born January 1, 2017 - March 15, 2019; Pandemic cohort, born November 15, 2019 and later

[b] Recommended age is 12 months; Pre-pandemic cohort, born January 1, 2017 - September 15, 2018; Pandemic cohort, born March 15, 2019 and later

[c] Recommended age is 12 months; Pre-pandemic cohort, born January 1, 2017 - March 15, 2018; Pandemic cohort, born March 15, 2019 and later

[d] Recommended age is 18 months; Pre-pandemic cohort, born January 1, 2017 - March 15, 2018; Pandemic cohort, born September 15, 2018 and later

Table S3 **Vaccine uptake (%) of routine childhood vaccines during the pandemic (data ending November 30, 2021).**

| **Polio** | **Dose 2 uptake at 12 months**[a] |
|---|---|
| | **Intermediate** |
| Overall | 87 |
| Gender | |
| Male | 86 |
| Female | 87 |
| Income quintile | |
| Q1 (lowest) | 82 |
| Q2 | 87 |
| Q3 | 89 |
| Q4 | 85 |
| Q5 (highest) | 92 |
| Unknown | 90 |
| Residence | |
| Rural | 79 |
| Urban | 93 |
| Unknown | 76 |
| Winnipeg region of residence | |
| Northern suburbs | 93 |
| Inner city | 88 |
| Southern suburbs | 94 |
| Regional health authority of residence | |
| Winnipeg | 93 |
| Interlake-Eastern | 84 |
| Northern | 83 |
| Southern | 72 |
| Prairie Mountain | 88 |
| Public Trustee / In CFS care | 76 |
| **DTaP** | **Dose 3 uptake at 12 months**[b] |
| | **Intermediate** |
| Overall | 77 |
| Gender | |
| Male | 77 |

| | |
|---|---|
| Female | 78 |
| Income quintile | |
| Q1 (lowest) | 67 |
| Q2 | 77 |
| Q3 | 81 |
| Q4 | 79 |
| Q5 (highest) | 86 |
| Unknown | 85 |
| Residence | |
| Rural | 69 |
| Urban | 84 |
| Unknown | 58 |
| Winnipeg region of residence | |
| Northern suburbs | 86 |
| Inner city | 78 |
| Southern suburbs | 87 |
| Regional health authority of residence | |
| Winnipeg | 85 |
| Interlake-Eastern | 75 |
| Northern | 62 |
| Southern | 66 |
| Prairie Mountain | 78 |
| Public Trustee / In CFS care | 58 |

| **MMR** | **Dose 1 uptake at 18 months[c]** |
|---|---|
| | **Intermediate** |
| Overall | 81 |
| Gender | |
| Male | 81 |
| Female | 81 |
| Income quintile | |
| Q1 (lowest) | 73 |
| Q2 | 82 |
| Q3 | 84 |
| Q4 | 84 |
| Q5 (highest) | 86 |
| Unknown | 81 |
| Residence | |

| | |
|---|---|
| Rural | 73 |
| Urban | 88 |
| Unknown | 82 |
| Winnipeg region of residence | |
| Northern suburbs | 90 |
| Inner city | 80 |
| Southern suburbs | 89 |
| Regional health authority of residence | |
| Winnipeg | 88 |
| Interlake-Eastern | 80 |
| Northern | 71 |
| Southern | 66 |
| Prairie Mountain | 84 |
| Public Trustee / In CFS care | 82 |

| **PCV** | **Dose 3 uptake at 18 months[c]** |
|---|---|
| | **Intermediate** |
| Overall | 77 |
| Gender | |
| Male | 78 |
| Female | 76 |
| Income quintile | |
| Q1 (lowest) | 70 |
| Q2 | 77 |
| Q3 | 80 |
| Q4 | 81 |
| Q5 (highest) | 81 |
| Unknown | 76 |
| Residence | |
| Rural | 69 |
| Urban | 83 |
| Unknown | 82 |
| Winnipeg region of residence | |
| Northern suburbs | 85 |
| Inner city | 77 |
| Southern suburbs | 85 |
| Regional health authority of residence | |
| Winnipeg | 84 |

| | |
|---|---|
| Interlake-Eastern | 77 |
| Northern | 65 |
| Southern | 63 |
| Prairie Mountain | 80 |
| Public Trustee / In CFS care | 82 |

| **MenC-C** | **Dose 1 uptake at 24 months[d]** |
|---|---|
| | **Intermediate** |
| Overall | 85 |
| Gender | |
| Male | 85 |
| Female | 84 |
| Income quintile | |
| Q1 (lowest) | 80 |
| Q2 | 85 |
| Q3 | 87 |
| Q4 | 85 |
| Q5 (highest) | 88 |
| Unknown | 84 |
| Residence | |
| Rural | 77 |
| Urban | 91 |
| Unknown | 87 |
| Winnipeg region of residence | |
| Northern suburbs | 92 |
| Inner city | 86 |
| Southern suburbs | 92 |
| Regional health authority of residence | |
| Winnipeg | 91 |
| Interlake-Eastern | 82 |
| Northern | 80 |
| Southern | 68 |
| Prairie Mountain | 87 |
| Public Trustee / In CFS care | 87 |

| **MMR** | **Dose 1 uptake at 24 months[d]** |
|---|---|
| | **Intermediate** |
| Overall | 85 |
| Gender | |

| | |
|---|---|
| Male | 85 |
| Female | 85 |
| Income quintile | |
| Q1 (lowest) | 80 |
| Q2 | 85 |
| Q3 | 87 |
| Q4 | 85 |
| Q5 (highest) | 89 |
| Unknown | 85 |
| Residence | |
| Rural | 77 |
| Urban | 91 |
| Unknown | 87 |
| Winnipeg region of residence | |
| Northern suburbs | 92 |
| Inner city | 86 |
| Southern suburbs | 92 |
| Regional health authority of residence | |
| Winnipeg | 91 |
| Interlake-Eastern | 83 |
| Northern | 80 |
| Southern | 69 |
| Prairie Mountain | 88 |
| Public Trustee / In CFS care | 87 |

| **Varicella** | **Dose 1 uptake at 24 months[d]** |
|---|---|
| | **Intermediate** |
| Overall | 84 |
| Gender | |
| Male | 84 |
| Female | 84 |
| Income quintile | |
| Q1 (lowest) | 80 |
| Q2 | 85 |
| Q3 | 86 |
| Q4 | 84 |
| Q5 (highest) | 88 |
| Unknown | 85 |

| | |
|---|---|
| Residence | |
| Rural | 77 |
| Urban | 90 |
| Unknown | 87 |
| Winnipeg region of residence | |
| Northern suburbs | 92 |
| Inner city | 86 |
| Southern suburbs | 91 |
| Regional health authority of residence | |
| Winnipeg | 90 |
| Interlake-Eastern | 82 |
| Northern | 80 |
| Southern | 68 |
| Prairie Mountain | 87 |
| Public Trustee / In CFS care | 87 |

| **Polio** | **Dose 3 uptake at 24 months[e]** |
|---|---|
| | **Intermediate** |
| Overall | 86 |
| Gender | |
| Male | 87 |
| Female | 86 |
| Income quintile | |
| Q1 (lowest) | 82 |
| Q2 | 87 |
| Q3 | 88 |
| Q4 | 86 |
| Q5 (highest) | 91 |
| Unknown | 90 |
| Residence | |
| Rural | 78 |
| Urban | 93 |
| Unknown | 83 |
| Winnipeg region of residence | |
| Northern suburbs | 94 |
| Inner city | 89 |
| Southern suburbs | 94 |
| Regional health authority of residence | |

| | |
|---|---|
| Winnipeg | 93 |
| Interlake-Eastern | 84 |
| Northern | 81 |
| Southern | 70 |
| Prairie Mountain | 88 |
| Public Trustee / In CFS care | 83 |

| **DTaP** | **Dose 4 uptake at 24 months[e]** |
|---|---|
| | **Intermediate** |
| Overall | 69 |
| Gender | |
| Male | 70 |
| Female | 68 |
| Income quintile | |
| Q1 (lowest) | 56 |
| Q2 | 70 |
| Q3 | 74 |
| Q4 | 74 |
| Q5 (highest) | 78 |
| Unknown | 72 |
| Residence | |
| Rural | 59 |
| Urban | 77 |
| Unknown | 50 |
| Winnipeg region of residence | |
| Northern suburbs | 79 |
| Inner city | 68 |
| Southern suburbs | 80 |
| Regional health authority of residence | |
| Winnipeg | 77 |
| Interlake-Eastern | 64 |
| Northern | 51 |
| Southern | 55 |
| Prairie Mountain | 75 |
| Public Trustee / In CFS care | 50 |

| **HiB** | **Dose 4 uptake at 24 months[e]** |
|---|---|
| | **Intermediate** |
| Overall | 69 |

| | |
|---|---|
| Gender | |
| Male | 69 |
| Female | 68 |
| Income quintile | |
| Q1 (lowest) | 56 |
| Q2 | 69 |
| Q3 | 74 |
| Q4 | 73 |
| Q5 (highest) | 77 |
| Unknown | 71 |
| Residence | |
| Rural | 59 |
| Urban | 77 |
| Unknown | 50 |
| Winnipeg region of residence | |
| Northern suburbs | 79 |
| Inner city | 68 |
| Southern suburbs | 79 |
| Regional health authority of residence | |
| Winnipeg | 77 |
| Interlake-Eastern | 63 |
| Northern | 51 |
| Southern | 54 |
| Prairie Mountain | 74 |
| Public Trustee / In CFS care | 50 |

DTaP = Diphtheria, tetanus, and acellular pertussis vaccine

MMR = Measles, mumps, and rubella vaccine

PCV = 13-valent pneumococcal conjugate vaccine

MenC-C = Meningococcal conjugate serogroup C vaccine

HiB =Haemophilus influenzae type b vaccine

[a] Recommended age is 4 months; Intermediate cohort, born March 15, 2019 - November 15, 2019

[b] Recommended age is 6 months; Intermediate cohort, born March 15, 2019 - September 15, 2019

[c] Recommended age is 12 months; Intermediate cohort, born September 15, 2018 - March 15, 2019

[d] Recommended age is 12 months; Intermediate cohort, born March 15, 2018 - March 15, 2019

[e] Recommended age is 18 months; Intermediate cohort, born March 15, 2018 - September 15, 2018

Table S4 **Changes in vaccine uptake (%) of routine childhood vaccines during the pandemic according to region of residence and income quintile (data ending November 30, 2021).**

| DTaP | Dose 3 uptake at 12 months[a] | | | | Dose 4 uptake at 24 months[b] | | | |
|---|---|---|---|---|---|---|---|---|
| | Pre-pandemic | Pandemic | Relative change | Fold change | Pre-pandemic | Pandemic | Relative change | Fold change |
| Overall | 80 | 74 | -7% | | 71 | 64 | -10% | |
| Winnipeg | | | | | | | | |
| Q1 (lowest) | 81 | 77 | -5% | -47.6 | 70 | 62 | -12% | 4.7 |
| Q2 | 87 | 85 | -3% | -25.2 | 79 | 72 | -9% | 3.5 |
| Q3 | 89 | 87 | -3% | -25.4 | 81 | 79 | -4% | 1.4 |
| Q4 | 91 | 88 | -3% | -33.7 | 84 | 80 | -4% | 1.8 |
| Q5 (highest) | 91 | 91 | 0% | ref. | 85 | 83 | -2% | ref. |
| Interlake-Eastern | | | | | | | | |
| Q1 (lowest) | 70 | 45 | -36% | 3.6 | 63 | 40 | -36% | 37.8 |
| Q2 | 84 | 77 | -8% | 0.8 | 71 | 68 | -4% | 4.1 |
| Q3 | 78 | 68 | -13% | 1.3 | 61 | 58 | -5% | 5.7 |
| Q4 | 81 | 77 | -4% | 0.4 | 70 | 70 | 0% | -0.1 |
| Q5 (highest) | 84 | 75 | -10% | ref. | 77 | 77 | -1% | ref. |
| Northern | | | | | | | | |
| Q1 (lowest) | 51 | 40 | -21% | 4.1 | 39 | 30 | -23% | 13.6 |
| Q2 | 63 | 41 | -34% | 6.7 | 51 | 30 | -41% | 23.9 |
| Q3 | 68 | 56 | -18% | 3.5 | 58 | 40 | -31% | 18.1 |
| Q4 | 80 | 71 | -12% | 2.3 | 67 | 54 | -20% | 11.9 |
| Q5 (highest) | 88 | 84 | -5% | ref. | 75 | 73 | -2% | ref. |
| Southern | | | | | | | | |
| Q1 (lowest) | 55 | 43 | -23% | 5.7 | 44 | 33 | -25% | 2.2 |
| Q2 | 66 | 55 | -17% | 4.3 | 61 | 49 | -20% | 1.8 |
| Q3 | 67 | 62 | -8% | 1.9 | 61 | 54 | -12% | 1.0 |
| Q4 | 66 | 56 | -15% | 3.6 | 61 | 51 | -17% | 1.6 |
| Q5 (highest) | 73 | 70 | -4% | ref. | 68 | 61 | -11% | ref. |

| | | | | | | | | |
|---|---|---|---|---|---|---|---|---|
| Prairie Mountain | | | | | | | | |
| Q1 (lowest) | 73 | 65 | -11% | 3.5 | 64 | 53 | -17% | 1.1 |
| Q2 | 83 | 77 | -8% | 2.4 | 74 | 67 | -9% | 0.6 |
| Q3 | 88 | 84 | -5% | 1.5 | 80 | 76 | -5% | 0.3 |
| Q4 | 88 | 84 | -5% | 1.4 | 80 | 76 | -5% | 0.3 |
| Q5 (highest) | 85 | 82 | -3% | ref. | 84 | 71 | -16% | ref. |

| MMR | **Dose 1 uptake at 18 months[c]** | | | | **Dose 1 uptake at 24 months[d]** | | | |
|---|---|---|---|---|---|---|---|---|
| | **Pre-pandemic** | **Pandemic** | **Relative change** | **Fold change** | **Pre-pandemic** | **Pandemic** | **Relative change** | **Fold change** |
| Overall | 82 | 75 | -9% | | 86 | 80 | -7% | |
| Winnipeg | | | | | | | | |
| Q1 (lowest) | 83 | 77 | -7% | 2.2 | 88 | 83 | -6% | 4.8 |
| Q2 | 89 | 85 | -5% | 1.5 | 92 | 89 | -4% | 3.5 |
| Q3 | 90 | 88 | -2% | 0.6 | 92 | 92 | -1% | 0.6 |
| Q4 | 92 | 89 | -3% | 0.9 | 93 | 91 | -2% | 1.8 |
| Q5 (highest) | 93 | 90 | -3% | ref. | 94 | 93 | -1% | ref. |
| Interlake-Eastern | | | | | | | | |
| Q1 (lowest) | 76 | 57 | -25% | 7.7 | 82 | 67 | -19% | -13.4 |
| Q2 | 81 | 74 | -9% | 2.6 | 82 | 77 | -6% | -4.4 |
| Q3 | 77 | 70 | -8% | 2.5 | 81 | 80 | -1% | -0.9 |
| Q4 | 81 | 77 | -5% | 1.5 | 84 | 84 | 0% | 0.3 |
| Q5 (highest) | 85 | 82 | -3% | ref. | 86 | 87 | 1% | ref. |
| Northern | | | | | | | | |
| Q1 (lowest) | 68 | 53 | -22% | 3.9 | 79 | 65 | -18% | 16.0 |
| Q2 | 77 | 52 | -33% | 5.8 | 89 | 71 | -19% | 17.3 |
| Q3 | 78 | 55 | -30% | 5.3 | 86 | 74 | -13% | 11.9 |
| Q4 | 82 | 61 | -26% | 4.5 | 92 | 70 | -24% | 21.4 |
| Q5 (highest) | 88 | 83 | -6% | ref. | 88 | 87 | -1% | ref. |
| Southern | | | | | | | | |
| Q1 (lowest) | 59 | 45 | -23% | 3.8 | 66 | 51 | -22% | 3.0 |
| Q2 | 69 | 58 | -16% | 2.5 | 74 | 61 | -17% | 2.2 |
| Q3 | 68 | 62 | -9% | 1.4 | 73 | 68 | -7% | 0.9 |

| | | | | | | | | |
|---|---|---|---|---|---|---|---|---|
| Q4 | 67 | 57 | -15% | 2.4 | 72 | 60 | -16% | 2.2 |
| Q5 (highest) | 76 | 71 | -6% | ref. | 78 | 72 | -8% | ref. |
| Prairie Mountain | | | | | | | | |
| Q1 (lowest) | 78 | 67 | -15% | 1.7 | 85 | 72 | -15% | 2.5 |
| Q2 | 84 | 73 | -12% | 1.4 | 86 | 76 | -12% | 2.0 |
| Q3 | 87 | 81 | -7% | 0.7 | 90 | 85 | -5% | 0.8 |
| Q4 | 88 | 83 | -6% | 0.6 | 92 | 87 | -5% | 0.9 |
| Q5 (highest) | 90 | 82 | -9% | ref. | 93 | 88 | -6% | ref. |

DTaP = Diphtheria, tetanus, and acellular pertussis vaccine

MMR = Measles, mumps, and rubella vaccine

[a] Recommended age is 6 months; Pre-pandemic cohort, born January 1, 2017 - March 15, 2019; Pandemic cohort, born September 15, 2019 and later

[b] Recommended age is 18 months; Pre-pandemic cohort, born January 1, 2017 - March 15, 2018; Pandemic cohort, born September 15, 2018 and later

[c] Recommended age is 12 months; Pre-pandemic cohort, born January 1, 2017 - September 15, 2018; Pandemic cohort, born March 15, 2019 and later

[d] Recommended age is 12 months; Pre-pandemic cohort, born January 1, 2017 - March 15, 2018; Pandemic cohort, born March 15, 2019 and later
